# “My doctor self and my human self”: A qualitative study of physicians’ presentation of self on social media

**DOI:** 10.1101/2023.09.27.23296214

**Authors:** Lauren A. Maggio, Lucía Céspedes, Alice Fleerackers, Regina Royan

**Author notes:** Correspondence should be addressed to Lauren A. Maggio, Department of Medicine, Uniformed Services University of the Health Sciences, 4301 Jones Bridge Rd., Bethesda, MD 20814. @LaurenMaggio.

## Abstract

**Introduction:** When using social media, physicians are encouraged and trained to maintain separate professional and personal identities. However, this separation is difficult and even undesirable, as the blurring of personal and professional online presence can influence patient trust. Thus, to develop policies and educational resources that are more responsive to the blurring of personal and professional boundaries on social media, this study aims to provide an understanding of how physicians present themselves holistically online.

**Methods:** 28 physicians based in the United States that use social media were interviewed. Participants were asked to describe how and why they use social media, specifically Twitter (rebranded as “X” in July 2023), which is especially popular among physicians. Interviews were complimented by data from participants’ Twitter profiles. Data were analyzed using reflexive thematic analysis guided by Goffman’s theory of presentation of self. This theory uses the metaphor of a stage to characterize how individuals attempt to control the aspects of the identities—or *faces—*they display during social interactions.

**Results:** We identified seven faces presented by the participants. Participants crafted and maintained these faces through discursive choices in their tweets and profiles, which were motivated by their perceived audience. We identified overlaps and tensions that arise at the intersections of faces, which posed professional and personal challenges for participants.

**Conclusions:** Physicians strategically emphasize their more professional or personal faces according to their objectives and motivations in different communicative situations, and tailor their language and content to better reach their target audiences. While tensions arise in between these faces, physicians still prefer to project a rounded, integral image of themselves on social media. This suggests a need to reconsider social media policies and related educational initiatives to better align with the realities of these digital environments.

## Introduction

Physicians increasingly use social media for multiple reasons, ranging from professional development^1^ to providing timely, accurate health information to the public.^2–4^ This online engagement can become a component of physicians’ *professional identity*^5,6^—their “representation of self, achieved in stages over time, during which the characteristics, value, and norms of the medical profession are internalized, resulting in individual thinking, acting and feeling like a physician”.^7^

Physicians and medical students are encouraged and trained to maintain and present separate personal and professional identities on social media.^8–10^ Yet, maintaining this separation is not always possible in practice, as physicians and medical students note that “doctors can’t be doctors all of the time” in these digital spaces.^11^ Specifically, the public nature of many social media platforms can make it challenging for users to control the content that is shared about them and to whom it is visible (e.g., a friend may post a photo of a physician depicting irresponsible behavior without asking for permission, which may then be viewed by colleagues or patients).^12,13^ Such *context collision* has created an “online identity crisis” for physicians, with negative implications for their well being,^14^ their professional reputation,^15^ and, potentially, the public’s trust in them.^13^

At the same time, the blurring of personal and professional identities that can take place on social media has the potential to benefit the patient-physician relationship when done with care,^14^ as experts who appear warm and personable are also perceived as more trustworthy, ^16,17^ and physician narratives have been shown to improve the perceived effectiveness of public health recommendations on social media.^18^ Yet, despite these potential benefits and the “operationally impossible” task of preventing context collision entirely,^14^ most existing policies and educational initiatives encourage physicians to practice complete professionalism on social media.^19–21^ To develop policies and educational resources that are more responsive to the blurring of boundaries that is inescapable online and guide physicians in navigating it effectively, medical educators must first understand how physicians present themselves on social media. This is particularly relevant as physicians with minoritized identities more frequently face harassment on social media, and report changing the way they use social media due to this harassment.^22^

Bearing in mind that social media platforms are not a “neutral stage of self-performance - they are powerful tools for consciously reconstructing multiple selves”,^23^ this study aims to describe physicians’ presentation of self on Twitter, including the aspects of their personal and professional identities they choose to present. We focus on Twitter, as it is one of the most popular with physicians^24^ and provides opportunities for dynamic, interactive identity presentation.^25^ Specifically, it allows users to post and respond to short messages that are freely available on the platform, enabling them to share multiple aspects of their personalities to a potentially broad audience.^12^

To describe physicians’ presentation of themselves we selected Irving Goffman’s theory of presentation of self, which uses the metaphor of a stage to characterize how individuals attempt to control the aspects of their identities—or *faces—*they display during social interactions.^26^ In this way, individuals “perform on multiple stages, creating a face for each interaction and developing faces for a variety of situational contexts”.^27^ The faces individuals display, and the *impression management* strategies they use to maintain them, are shaped by their beliefs about the audiences who are watching and what those audiences expect, as well as the feedback they receive in response to their ‘performances.’

Although Goffman’s theory was originally conceived in relation to face-to-face interactions, it has been found relevant for describing identities in social media contexts,^28^ including among physicians.^3^ In addition, researchers have expanded the theory beyond its original focus on the active performance of faces to also consider static elements (e.g., photos, curated lists, etc.) that individuals curate to leave a particular impression on their audiences.^29^ This paper considers both participants’ performances (i.e., their Twitter activities) and exhibitions (i.e., their Twitter bio statement and images) to provide a more holistic picture of physicians’ online self-presentation.

## Methods

We conducted an interview study guided by a constructivist paradigm, which means we believe that knowledge is socially constructed between individuals (e.g., participants, researchers) and society.^30^ Iterative engagement with participants’ Twitter profiles and descriptions of their social media was used to co-construct an understanding of how physicians present their identities on Twitter. By taking this constructivist approach, our backgrounds and experiences informed this study, including data interpretation and analysis. To provide multiple viewpoints on the topic, the research team included expertise in science communication (LC), health communication (AF), health professions education and scholarly communication (LM), and clinical medicine and social media (RR). RR is also an emergency medicine physician knowledgeable about and active on social media and is a member of the Illinois Medical Professionals Action Collaborative Team (IMPACT), which leverages social media to combat misinformation.^2^ The Uniformed Services University ethics board declared this study exempt from further review (Case # DBS.2022.427).

Data was collected between October 2022–May 2023, and analyzed from November 2022– August 2023. During the study period, Twitter was a key platform for sharing credible, COVID-19-related information—including by physicians^31,32^—but also for circulating pandemic-related disinformation.^33–35^ In addition, the platform was purchased by Elon Musk in October 2022,^36^ which resulted in changes to Twitter’s blue check verification system (April 2023) and the rebranding of the platform as “X” (July 2023).

### Participants and Recruitment

Physicians who used social media were eligible to participate. We focused on US-based physicians so that participants would share a similar healthcare landscape. Initial recruitment was targeted towards physician members of the Association for Healthcare Social Media (AHSM) and IMPACT. Participants were recruited by email and direct messages on Twitter. We used snowball sampling^37^ to identify additional physicians active on Twitter by asking participants for suggestions during the interviews.

### Data collection

Data were collected using one-on-one interviews and supplemented with information from each participant’s Twitter profile (e.g., hashtags, photos). On May 3, 2023, each participant’s number of followers and tweets; geographic location; account start date; and bio statement was recorded.

Interviews were guided by a semi-structured interview guide based on a review of the literature and shaped by Goffman’s theory of presentation of self.^26^ We piloted the interview guide with two physicians who provided feedback on the content and structure, and then slightly modified the interview guide. The interview guide also evolved over the study based on researcher conversations and events related to Twitter (e.g., Musk’s purchase of the platform) Although we designed our initial interview guide to capture how and why participants use social media in general, our focus eventually shifted to Twitter, as this was the platform participants used most. (Appendix A).

Interviews lasted between 40-60 minutes on average and were conducted by LM and LC using Zoom or Google Meet. Zoom interviews were transcribed by a transcription service and Google Meet interviews via its built-in transcription feature. All interviews were deidentified.

Throughout the interviews, LM and LC met bi-weekly to discuss the transcripts, identify relevant codes, modify the interview guide as needed, and determine whether additional participant recruitment was required. The decision to end data collection was based on *information sufficiency*, the point at which we felt that we had captured enough data to answer our research questions.^38,39^ Based on our ongoing conversations, we determined that we had reached data sufficiency after interviewing 24 participants. However, we interviewed four additional participants to be certain, yielding a final sample of 28 participants.

### Analysis

We analyzed the transcripts using reflexive thematic analysis^40,41^ due to its alignment with our constructivist paradigm and its flexible nature and usefulness in facilitating the identification, examination, and reporting of patterns.^42^ Analysis was guided by Braun and Clarke’s six steps of thematic analysis.^40^ We began by independently reading the transcripts and bio statements line-by-line, then generated initial codes. While coding, we were sensitized to seek content aligned with Goffman’s theory, specifically the presence of “faces,” but also remained open to other codes. Codes were compiled into a working codebook that included preliminary code definitions and example quotes. We met frequently to discuss, reflect on, and refine the codebook. Once the codebook was agreed upon, LM and LC independently applied the codes across the transcripts using Dedoose, a qualitative software. Generated themes were shared with RR and AF for discussion, refinement, and naming. To verify the trustworthiness of our findings, member checking was conducted^43^ by presenting participants a summary of results and asking them if the findings resonated with their experiences.

## Results

The 28 physicians interviewed (n=18 women) represent 10 clinical specialties with pediatrics and emergency medicine being the most common. While all participants hold MD or DO degrees, five also have master’s degrees (e.g., in public health, business administration) and one a PhD. Twenty participants hold appointments at academic health centers.

All participants have Twitter accounts and 25 have a presence on additional platforms (e.g., TikTok, Facebook). Participants have a total of 646,328 Twitter followers (Mean=23,083), with individual follower counts ranging from 518 to 115,800. Together participants tweeted 1,054,660 times (Mean=37,666.4), with the number of tweets per participant ranging from 296 to 286,600 tweets over the lifetime of their accounts. Before Twitter transitioned to a subscription-based verification system in April 2023, 15 participants had verified profiles (i.e., featuring the blue checkmark); only a single participant retained this status post transition.

Participants present seven faces (See Table 1 for a listing of faces), which resonate with participants who responded to our member checking request. All participants engage in impression management, which is the process of attempting to influence how others view an individual. For our participants, impression management depends on three related factors across the faces: language choices, motivations, and audiences.

**Table 1.** Faces identified in physician interviews (n=28) with sample quotes.

| Face | Sample quotes |
| --- | --- |
| **Physician** | A lot of my professional identity is being a pediatric physician and caring for children, and so that is a big part of where my identity lies, being a pediatric physician. And so I work really hard to highlight and advocate for children and that, I think, is a reflection of my professional and personal identity on the Internet or on social media. (P26)<br>I think without listing MD I felt like you wouldn't necessarily know I'm a physician, so that's sort of why I put it there [Twitter profile]. (P20) |
| **Academician** | Initially when I was doing it [tweeting], I was actually doing it to amplify my own work and my own research and any publications that I had, because that was what had been recommended to me. (P2)<br>Twitter has given me a lot of opportunities that I wouldn't have otherwise had in terms of connecting with other researchers or people that are still serving as mentors today" (P1). |
| **Networker** | People have contacted me asking if I want to write a paper or even for jobs and things like that. Then I usually point them to other sources or boost other people. And like I mentioned, reporters have contacted me and those kinds of things. So it definitely helps. Just expand your range within your professional community and beyond that. (P14)<br>So, I've interacted with various legislators on the state and federal level and use it as a way to help tag what issues are important in the pediatric community. [...] There's definitely been other legislators who've reached out to me or who I've reached out to and was able to secure a meeting with or probably reach them in a different way, when calling or emailing them didn't work. (P7) |
| **Advocate** | It's a form of advocacy for me...I want to also share the message of climate change and how dire it really is and that people don't realize how bad it is, and that I want to – without taking away hope, but still conveying that this is actually something that's worth looking into, doing something about. (P4).<br>I also use it as an opportunity to highlight the challenges of gender equity and medicine. The challenges of just any sort of identity equity in medicine. There are a lot of underrepresented people in medicine, or undervalued people in medicine, and I try to use it as an avenue to showcase how to increase value or how to highlight the value of certain groups of people. (P26) |
| **Public educator** | One of the things that I try to do with my account is counter any misinformation that I see and put forward a perspective that's grounded in my own expertise. (P13)<br>I find that from a medical standpoint if I'm able to influence people's health decisions, I think I've made a huge mark on the world [...] if I can make a dent in that as one person I think I've made an even broader impact than just being a private practice physician. (P9) |
| **Human** | This is me. I'm a physician. I'm a woman. I'm a mother. (P27)<br>I feel like I should feel free to be myself as well. Because I feel like we are raising a generation of physicians who are terrified to have a life outside of medicine to be a person and that is what leads to such high rates of burnout, anxiety, suicide, among healthcare professionals and I refuse to contribute to that. And so I try very hard on what I include both in my profile, and in what I post to be a very real person to share the triumphs and hardships in medicine, but to share life outside of medicine. Because I am setting an example, for all the people that are following me, that life is not |
|  | medicine. Medicine is a job, it's an amazing one, but it is not my life... mainly on my profile and I think that it's a blend of saying, Yeah, I'm a doctor and that's important to me and that's also reflected in the content that I post, but I'm a person and you can be a person too, and that's why I've chosen the pictures and the information that I have featured. (P19) |

Participants construct and maintain these faces through discursive choices in their tweets and profiles, which are closely related to their motivations for engaging with different audiences on Twitter. In addition, participants are keenly aware of the heterogeneity of their potential audiences and work hard to project faces that fit the image they hope to convey to the audiences they believe are watching their ‘performance.’ One participant describes: “I am deliberate in the language that I choose depending on who the audience is” (P5). When describing their imagined audience, one participant says: “Honestly, I think it’s the world. I mean, anything that you put out there, anybody can see it” (P22).

We now describe the seven faces, as well as the overlaps and tensions that arise at their intersections. For each face identified, we highlight how participants describe their motivations, consider their imagined audiences, and select their language to leave a particular impression (Table 2).

**Table 2.** Faces identified in physician (n=28) interviews as aligned with their intended audience, motivation and examples of language choices.

| Face | Imagined audience | Motivations | Language Selection Examples |
| --- | --- | --- | --- |
| **Physician** | Colleagues and peers; patients; general public; employers and institutions | Asserting legitimacy within the medical professional community; establishing expertise and professionalism; showing compliance with good / professional / legal social media practices | I use my profession as a lens for how I decide what to post and the ideas I share. (P4)<br>Whenever I'm at the urge to post something, the first thing I think about is, is this professional? It doesn't violate any rule. I'm not going to post something nasty, I'm not gonna post something that would hurt a patient or another person intentionally. I'm not gonna post something that's gonna violate HIPAA. (P19) |
| **Academician** | Colleagues and peers | Promoting scientific research and scholarly achievements | If I'm tweeting out to a technical audience, then I would probably choose something that is, you know, more at their speed of technical paper. (P11)<br>I also use it [Twitter] just to share my own work and as an opportunity to collaborate with other people that might be in similar subjects. So when I see articles that are important and relevant that I think that other colleagues of mine might want to see, I'll retweet, re-share. (P18) |
| **Networker** | Colleagues and peers; journalists, policy makers, and advocacy groups | Creating connections for professional opportunities and academic collaborations | I think networking on Twitter plays out by connecting people, just like you connect people in real life. You can connect them by, you know, highlighting them in your posts or things like sharing the work, creating collaboration. (P26)<br>My bio statement I've changed a few times, and I honestly can't right now even recall what it says, but I've changed that a few times to sort of reflect the scope of my interests, knowing that Twitter is a facilitator for networking too, and I've engaged with a lot of folks through that, knowing you never know who's gonna look at your profile, and that's like a great way to convey what your interests are, that might not be reflected in your tweets or retweets. (P20) |
| **Advocate** | General public; policy and decision makers; journalists | Advancing and engaging with social causes | So after meeting in DC with the American Academy of Pediatrics Legislative conference, I started looking at using Twitter, seriously, as a form of connecting with other advocates. So, through their registered hashtag I was able to form a lot of connections with them and really work with the whole scope of pediatricians trying to advocate for similar things for our patients. (P7)<br>We kind of use this expression in advocacy, "the data writes the policy but anecdotes get the votes". And so, you know, if you can state the fact and say, you know, in my experience, I've seen<br>this as well or I will never forget the case of whatever that can be helpful. (P11) |
| **Public educator** | Patients; general public | Educating the public on health issues by sharing reliable information | We totally change how we talk, the words we use, the amount of words we use, you know, not the repeat signs of the people, but explaining complex medical things to people with understanding versus someone with no understanding of the context is a totally different ballgame. (P6)<br>In one message I might write about, you know, Ventricular fibrillation and how that can lead to being treated with the defibrillator that might be more for a scientific community versus when your heart stops, you know, you should do hands-only CPR and kind of describe that. So, I do change a little bit the language...because it's going to be different if I'm talking to a colleague about something that was published in a medical journal. Then I might just post the actual medical journal versus trying to get that information made to the public. Then I might use a news article, or a more lay piece, or maybe a blog post or something I think might be easily understood by the general public. (P18) |
| **Human** | Colleagues and peers; general public | Role modeling a relatable and grounded identity beyond the professional physician role | There's a great worry in the physician community about making sure you look professional and that patients can see us, and, you know, we're doctors, this is very serious. And yes of course but you can still be very professional and say "Look how cute my dog is in this picture". (P11)<br>What you see there is sort of an amalgamation of my doctor self and my human self. I post a lot about food and running and my kids. So I don't limit myself to just posting stuff as a physician and part of that is very much by design, to sort of demonstrate that physicians are human beings and have foibles like everyone else. And so I'm very intentional in sort of mixing my personal life with the fact that I'm a physician on Twitter." (P5) |

### Physician

All participants present a *physician face* in their online interactions: “when I interact with other people on Twitter, I do it as a physician” (P1). Most participants identify as an MD in their bio statement or handle (e.g., @ParticipantMD) and indicate their clinical specialty. Listing these credentials enables participants to establish themselves as legitimate members of the medical community and as reliable experts to patients and the public. Aware of this dual audience (i.e., colleagues/peers, patients/public), participants attempt to align their *physician face* with their professional identity and practices. One participant notes: “I’m using [Twitter] in a way that is congruent with who I am as a person but also what kind of physician I want to be”. (P7).

Publicly putting on a *physician face* online is not without risks. As Participant 22 states: “You can say 10,000 things right on social media. But if you say one wrong thing, it can end your career”. Therefore, physicians are motivated to don the *physician face* to present themselves as medical professionals who comply with the social media practices expected of a responsible physician, including self-imposed “rules of thumb” and explicit guidelines prescribed by employers or patient privacy laws.

The *physician face* considers participants’ relationships with their institution or organization, as well as their patients. It is visible in disclaimer language in their bio statements (e.g., “Tweets my own/not employers” (P7), “Tweets ≠ medical advice” (P21)) and in descriptions of impression management strategies from their interviews. For example, Participant 23 says they rely on their “Grandma Rule,” meaning they avoid posting anything they would not want their grandmother reading. Another notes: “I am mindful of the fact that a patient’s parent could see what I’m saying.” (P13).

The need to perform the *physician* face relates to internalized ideas of “how you should behave” (P26) on social media, which are shaped by a range of tacit and explicit rules and stances at participants’ institutions and professional associations. Participants at supportive institutions report experiencing positive recognition of their active social media presence, with some noting their institutions have even created specific social media positions for physicians. However, other institutions frown upon physicians tweeting, as Participant 19 describes: “I’ve been told that until I develop better, professional judgments, basically get off Twitter, I will not be promoted.” Physicians in private practice cite greater autonomy, underscoring the power of institutional context in participants’ self-presentation: “I have a certain degree of liberty to just sort of figure it out for myself and do it as I please, because I’m in private practice and I’m one of the partners in the practice. And so, I’m my own boss and I don’t have to worry about offending any higher-ups.” (P13).

This variability in institutional approaches causes a tension, leaving participants acutely aware of their responsibilities as physicians to both patients and employers. They must think carefully about the language, content, and tone of their tweets, to avoid ire from employers and patients. For many participants, these impression management strategies are also important for protecting their academic careers, such that the *physician face* becomes entangled with the *academician face*.

### Academician

Participants consider promotion of scholarship to be an extension of their academic and professional practices. They don their *academician face* in their Twitter profiles, often by including their academic role (e.g., “professor of pediatrics”) and a link to their institutional profile. When putting on this face, participants both produce and engage with biomedical scientific content with an intended audience of peer health professionals and scientists. Consequently, the language of their tweets tends to be more technical, including links to journal articles. Hashtags used (if any) are specific to the topic in question or the specialist circle addressed.

Participants are motivated to use the *academician face* to promote their own and other’s scholarship, with many describing this potential for promotion and, ultimately, academic success as what motivated them to join Twitter. As Participant 18 noted,

> […] everything is, let’s face it, currency for your self promotion. It’s largely based on how visible your public figure is. […] you might be a fantastic researcher. But if your only readers are going to be people that actually are subscribed to the journal that you wrote your publication to, then that’s not going to have the same impact. And so there is a little bit of serving and self-promotion in this and trying to make your brand visible.

Parallel to this focus on self promotion, some physicians use this face to promote medical students and residents (e.g., by sharing co-authored publications, tweeting trainee publications to boost visibility). Physicians describe this aspect of the *academician face* as an extension of their offline mentorship responsibilities: “I’m in the position where I don’t need that much help myself because I’m in a senior position but I feel, like, how to give back and that’s what I do. I mentor a lot of people at all different levels, starting from undergrad to medical students to residents” (P14). This mentoring aspect of the *academician face* relies heavily on projecting a positive disposition towards networking, both for professional gain (e.g., reaching potential collaborators) and social connection. The *academician face* therefore closely connects to the *networker face*.

### Networker

Participants are primarily motivated to use the *networker face* to connect with colleagues and peers in order to secure professional opportunities and collaborations. When successful, this networking is described as highly rewarding:

I think the connections that I’ve made through this sort of community on Twitter have definitely helped me professionally. I got an editorial fellowship that *I don’t know if I would have gotten otherwise*, and just have gotten other good opportunities that I feel pretty strongly came from those connections on Twitter. (P17, emphasis added).

We find no explicit descriptions of how participants tailor their language to perform this face. Yet participants do note the importance of projecting a certain disposition (e.g., as resourceful, approachable, friendly). In addition, participants describe actions such as retweeting and tagging and direct messaging specific individuals or professional associations as ways to display their openness to networking.

Interestingly, *refraining* from these actions can also be important for projecting a *networker face*, at least in the early ‘scenes’ of the performance. Many participants say they encourage physicians who are new to Twitter to spend time just “lurking” on the platform before engaging professionally with others. The *networker face*, then, is gradually built according to each individual’s interests, needs and motivations in different stages of their professional trajectories.

While colleagues and peers are the primary audiences of the *networker* face, physicians also perform this face with other social agents in mind, including journalists, policy makers, and advocacy groups. The *networker* face, therefore, can be a strategy for platform-building that blends into the *advocate face*.

### Advocate

Participants don their *advocate face* when advocating for social causes or “passions,” often through linguistic choices in their bio statements (e.g., hashtags such as #DiversityInMedicine or #WIM[Women In Medicine]StrongerTogether). Physicians deploy the *advocate face* when taking a public stand on issues characterized by some level of social disagreement or controversy (e.g., climate crisis, police violence), most often those that relate to social aspects of medicine (e.g., reproductive rights, access to healthcare):

> I’m not afraid to shy away from some of those opinions because I know a lot of people are wanting to know where I stand on those things […] they know that I’m actively invested in this endeavor to help educate people and also from a humanistic standpoint. I talk about social justice issues as well (P9).

Participants also advocate for change within medical professions, such as reducing race, gender, or class inequities among medical students or increasing diversity among physicians or researchers. Some also use Twitter as a channel for communicating their participation in activities such as protests and campaigns (e.g., a rally for gun control). Participants describe this advocacy as important to them on a personal level but also for the medical profession generally:

> I have advocated for disabled students. And it’s really very effective, it turns out, because you get a wide audience, and organizations are, in a way, forced to respond to the concerns you bring. You can really amplify the cause you’re pushing. Because of what I’ve tweeted or something, reporters for example have contacted me to do interviews (P14).

Since the *advocate face* often relates to contentious social issues, some participants describe this face as in tension with their *physician face*, especially in relation to their employer audience. One participant recollects that “a couple of times they’ve [the social media office] contacted me to say, ‘You know, the optics of this are not what we would prefer. If you would not mind, would you change the wording or take something down?’” (P7).

While other medical professionals are one possible audience of the *advocate face,* participants also engage society more broadly (e.g., promoting issues to the media, making a hashtag go viral), including interacting in a public forum for addressing and engaging with politicians. Participants describe the public aspects of their *advocate face* as having been particularly important during the COVID-19 pandemic, when combating mis/disinformation as a form of advocacy. Thus, the pandemic has created a situation in which physicians’ *advocate face* blurs with another face: that of the *public educator*.

### Public Educator

Participants are highly motivated to use Twitter to educate the public on health issues and thus put on the *public educator face*. This face is, in part, an extension of their daily patient interactions, as seen in comments such as, “I do think that my role has to extend beyond the bedside. That I should take advantage of the platform that I have to try to counteract misinformation and to provide good reliable information.” (P18). Thus, physicians tweet to share curated information that complements, expands, or fills in what they perceive to be gaps in patients’ understanding of medical topics. “A lot of the time I pick what I’m gonna write about or tweet about or make a video about according to what is coming up in the office. What is the most confusing thing for my patients?” (P21).

Yet, participants also recognize that social media significantly expands their audience to include virtually anyone. Thus, they perform the *public educator face* to educate their patients, but also to inform a wider public: “I feel like it’s part of my job. It’s part of what I signed up for as being a physician to educate my communities. And my communities are much larger than what they used to be.” (P2). Participants’ awareness and perceptions of these larger communities, in turn, shape how participants draft their tweets:

> If I’m going to tweet [to] the general population, I just keep in mind what kind of message I want to convey, what kind of tone I want to convey it in, what my goal is with that. […] I do use my credentials in there because I want people to know where this information is coming from, but I might use slightly different language talking to people who are not in the medical field versus the medical field. Just like I would in the hospital. (P7)

Many participants had experience communicating health information on Twitter before the pandemic. During COVID-19, these physicians capitalized on their expertise and connections to reach and, in many cases, expand their follower base, as they turned from sharing specialized or general medical educational content to disseminating quality information related to the new and unknown virus. While a thorough analysis of the *public educator* face during the pandemic is beyond this paper’s scope, participants describe some aspects of their performance that appear to be unique to the crisis context, including sharing graphical material (e.g., infographics); posting recommendations and practical information; and practicing special care when sharing claims about controversial topics (e.g., masking).

> I know if I post this, I’m gonna get like a million people saying terrible things about me, but then I kind of realized, well, is there benefit to that post? Is it an educational post? Is it gonna help somebody if it is? All right, then we’ll take one for the team […] At the end of the day, if we can still get good information out and somebody is seeing it and getting benefit from it, then keep doing it (P11).

Descriptions of personal experiences treating patients with COVID-19 were another common way in which participants performed the *public educator face* during the pandemic. In this sense, participants opened up online by showing their *human face*.

### Human

Physicians perform the *human face* by disclosing details about their personal identities. This includes tweeting about experiences of failure, emotions, or mental health; commenting on family life; describing their race and sexual orientation; or sharing light-hearted content on topics such as sports, music, or hobbies. Participants also display their *human face* visually, posting casual headshots as their profile pictures and nature photographs, memes, or cartoons as their background banners. Multiple participants include their pronouns in their bio statements, while others mention marital status or family members (e.g., “Dad of 4,” “Mom, wife”). Bios also describe interests or identity traits, including statements such as: “Loves baseball,” “Proud Iranian/American.”

Participants don their *human face* for fellow physicians and trainees to encourage a healthy approach to balancing personal and professional responsibilities. As P27 explains, “I want to role model that I’m a physician with a life. A physician with a life and roles and identity outside of work, and I can’t just turn one role off and turn on the other. They intersect day in and day out.” Participants also perform this face with patients and the wider public in mind with a goal of appearing more relatable:

> I think I get credibility for demonstrating humanity, honestly. I think it allows people to see me as me. And I think that once they sort of know who I am as a human, I like to think that it, I hope, makes them more likely to listen when I say something medically. (P5)

While participants believe performing the *human face* is important they acknowledge that doing so can create tension with the *physician face*. For example, P21 believes that, “it’s important to show your humanity” on Twitter, but notes that many trainees tweet about content that is “way more personal” than they would be comfortable sharing themselves (P21). Other participants describe experiencing employer pushback:

> It’s become more personal to me. I’m not gonna lie, that’s got me in trouble at work. Even when I don’t post things that are at all inappropriate, I’ve been told that it is unprofessional and shows poor judgment to post dog pictures several times a day because physicians should not be doing that. I’ve been told a physician should not admit that they have their own health problems online because patients wouldn’t want to see a doctor who’s not perfectly healthy. (P19)

Although participants note that presenting the *human* and *physician face* simultaneously can be challenging, they also see it as important: “I want [social media] to be like a true representation of who I actually am. I don’t want to create this persona that’s divorced from how I actually am” (P13). Perhaps for this reason, participants maintain only one Twitter account as opposed to separate professional or personal accounts. The *human* face thus recurs throughout participants’ profiles and content, incorporating an element of authenticity and relatability into the other faces they present:

> You paint yourself as a person, which I try to do, you know, as a dad, as a dog lover, as a photographer. And I think that grows you as a physician, as a practitioner, and it shows who you are to the community and to your patients and to your colleagues. It’s sort of an extension of who you are as a person (P22).

## Discussion

We use Gofffman’s theory of presentation of self to explore how physicians navigate the complexities of identity expression on Twitter—a platform that allows users to share multiple aspects of their personal and professional selves, but offers little control over who ultimately witnesses these acts of self-presentation.^12,25^ Participants overwhelmingly embrace this opportunity for dynamic self expression, performing multiple roles, or faces, simultaneously on the Twitter stage. Some of these faces, such as those of the *physician*, *academician*, and *networker*, are more closely tied to physicians’ professional roles—motivated by a need to assert medical legitimacy, promote scholarly achievements, or increase career opportunities. Others, such as the *public educator face,* expand physicians’ professional duties (i.e., to inform and educate) but expand the target group of those duties from patients to a wider, less defined, digital public. Still others, such as the *advocate* or *human face*, allow physicians to share more personal aspects of their identities, such as their passion for social causes, mental health struggles, and hobbies.

Physicians see the ability to openly display their more intimate experiences and characteristics as an advantage of Twitter, as it enables them to role model a healthy relationship to personal identity that they feel will benefit peers and trainees. This desire is understandable given medicine’s history of occupational distress,^44^ which is linked to physician burnout^45^ and suicide.^46^ This use of the Twitter stage also aligns with recent calls for new approaches to wellness that encourage physicians to blend their personal and professional lives and, ultimately, shift medical culture away from perfection and toward vulnerability and self-compassion.^47^ While these calls were not necessarily drafted with social media in mind, participants’ embrace of the personal on Twitter could potentially support this new approach and improve physicians’ quality of life.

While physicians in this study value sharing “an amalgamation of my doctor self and my human self,” (P27) they also acknowledge that tensions can arise as the boundaries between the professional and personal become blurred, which aligns with previous studies.^5,11,48^ We observe these tensions most often between the highly professional *physician face* and the more personal *human* and *advocate faces,* and note their close connection to the imagined audience(s) of each face. As physicians don their different faces, they strive to engage diverse audiences—including peers, trainees, journalists, policymakers, patients, and the general public—and carefully craft their tweets and profiles to portray the traits they think are most appropriate for each group. Simultaneously, they recognize that the nature of the digital ‘stage’ on which they are performing makes doing so impossible, as the publicness of the platform means audiences who were never intended to witness particular acts of self-presentation can easily end up with a front-row seat.^12,25^ Physicians develop impression management strategies to mitigate the negative consequences associated with such context collapse, for example, noting in their bios that their tweets reflect their own views only, and not their employer’s, or self-censoring content with the potential to violate HIPAA or harm patients. Yet despite their best efforts, considerable overlap between physicians’ faces nevertheless takes place. Social media guidelines recommend that physicians separate personal from professional expressions of identity as much as possible,^14^ but such separation is perceived as unrealistic—or, in many cases, even undesirable—for participants. This suggests a need to reconsider these ‘best’ practices for using social media to better align with the realities of these digital environments. Moreover, medical educators must ensure that educational initiatives embrace these realities and tailor training accordingly.

Revising social media guidelines to encourage—rather than prohibit—thoughtful expressions of nonprofessional identities could also prove beneficial for the public, as previous research has revealed that experts are more trusted when they are perceived as warm and personable^16,17^, and find public health messaging to be more effective when it includes physician personal narratives.^18^ Indeed, physicians in this study believe that exhibitions of humility and humanity— exemplified in the *human face*—can help patients and the public better relate to them. They take advantage of Twitter’s affordances to display this face, but also to inform the public and combat misinformation. This *public educator face* leads physicians to share reliable health evidence in ways that non-experts can use and understand and is seen as an extension of their offline professional responsibilities. The subjects in this study felt strongly about this role, despite concerning levels of harassment of physicians and scientists on social media.^22^ This commitment to public education via social media is encouraging, given that physicians remain one of the most trusted sources of health information among the public.^49^ As online health misinformation proliferates,^33^ physicians can take advantage of the increasing availability of social media platforms to inform and protect the public.

### Limitations

First, our study may be limited in generalizability outside of the US, only US physicians were included. Most participants are also affiliated with academic medical centers, which may have impacted our findings, especially in relation to the *academician and public educator faces*. This study also focuses on physicians’ self-presentation on Twitter—now “X”—a platform which is rapidly evolving. Paired with the pandemic context in which this research was conducted, this focus means that our findings may not apply to current realities on the platform, nor to other platforms or different time periods. Future research could thus build on our results by examining physician self-presentation on other platforms, during other crisis (or noncrisis) contexts, among other groups of physicians, and within other world regions.

## Conclusions

This study delves into physicians’ self-presentation on Twitter, and identifies seven faces they perform on Twitter. Each of these is motivated by different communicative objectives and requires a particular kind of impression management to be maintained. To bring each face to the stage, participants tailor the language and content of tweets to the specific audiences they imagine are watching.

Social media, and Twitter in particular, present physicians an opportunity to express elements of their identity that they are generally not encouraged to reveal in professional daily interactions. These findings reveal that physicians are aware of the potential risks posed by this more personal presentation of self, yet still find it more rewarding than reducing their use of social media to the maintenance of a purely professional face. By not detaching their personal and professional identities, but instead strategically choosing when to emphasize each of their faces, they challenge traditional representations of the way in which physicians should present and conduct themselves. “It’s a blend of saying, Yeah, I’m a doctor and that’s important to me and that’s also reflected in the content that I post, but I’m a person and you can be a person too.” (P19)

## Ethical Approval

This protocol was deemed exempt by the Human Research Protection Program at the Uniformed Services University of the Health Sciences (Case # DBS.2022.427).

## Data

None reported

## Disclaimer

The views expressed in this article are those of the authors and do not necessarily reflect the official policy or position of the Uniformed Services University of the Health Sciences, the U.S. Department of Defense, or the U.S. Government.

## Data Availability

All data produced in the present study are available upon reasonable request to the authors.

