## Appendix A for "“My doctor self and my human self”: A qualitative study of physicians’ presentation of self on social media"

**Interview Guide**

**Physician’s use of social media**

**General icebreaker question:**

To start off, please describe how you use social media as a physician.

**Impression Management/Professional Identity**

- In what ways, if any, do you attempt to manage the impression you make on social media?
  - Why do you think this type of management is necessary or important?
- What do you think your profile says about you?
  - How did you select the images for your account (e.g., your bio photo and background)?
  - How did you choose your bio statement and hashtags?
  - What made you select your pinned tweet?
  - How often do you modify your profile and what will prompt a modification?
- How would you describe your identity on social media?
  - In what ways does your identity on social media align with or extend your professional identity as a physician?
  - Vice versa, are there any ways that you feel your social media persona influences your physician identity?
  - If I were a patient that followed you on Twitter and I walked into your office, in what ways, if any, would your personality be different from social media?
    - What about if I were a medical student or resident?
    - What about if I were your colleague?
    - What if I were your department chair? (For academics)
- Can you provide an example of when your personal and professional identities converged on social media?
  - How did you handle it?
  - Has your visibility on social media ever caused problems for you?

**Audience**

- When you post, who do you imagine your audience to be?
  - In what ways, if any, do you tailor your approach to that imagined audience?
  - On social media researchers talk about two audiences, the one we intend to communicate with and then the one we subconsciously are trying not to offend. Who do you think you are trying not to offend and why?
  - What sorts of engagement do you have with your audience?
    - Can you describe an example of when this engagement has been demotivating and made you question your use of social media?
  - In what ways, if any, have your physician peers and/or institution reacted to your use of social media?

**Backstage**

- Some participants have described social media as a space where they can show the more human side of their physician identity. Does this resonate with your experience?
  - If yes, can you provide an example of how you have shown your “more human side” on social media? How did people react?
  - If no, why not?
- Do you engage with people on Twitter via DM?
  - If yes, who do you tend to engage with?
  - Can you give an example of a situation when you would shift from your public feed to communicating via DM's?
  - In what ways do you think might interact differently when utilizing DMs?

**Pandemic**

So now, I want to change gears and talk about the pandemic. During the pandemic, what did you feel was at stake for physicians engaging in social media? Credibility, trust, reputation?

- What do you think was at stake for you personally?

During the pandemic there was a lot of scientific uncertainty. In what ways, if any, did you attempt to provide clarity during this time on social media?

- How did you decide what information to post about and how did you evaluate this information?
- What did you find most challenging about communicating reliable science and health information in this context?
- In what ways, if any, did you put disclaimers on information you shared?
- Who did you look to to validate information during this time?
- How did you rely on the physician community?
- When posting COVID related information, did you perceive any changes in the feedback / comments / engagement you were used to prior to the pandemic?

People have described information shared during the pandemic as value-loaded. Does that resonate with you?

- Can you give me an example?
- How do you think this influences people’s trust in science?
- In what ways, if any, do you think you communicate your values in your social media posts?

How did the pandemic influence how you engage with social media?

- In what ways, if any, did the pandemic influence your motivation to post?
- For content related to COVID, did you take different things into consideration that you might not have if the content wasn’t about the pandemic?
- In what ways, if any, do you think your use of social media during the pandemic was negative?
- Do you feel the pandemic has changed your social media habits? If yes, do you feel you will keep these new attitudes/habits?

During the COVID-19 pandemic many physicians turned to social media. Why do you think this happened?

- Some physicians we have spoken with have indicated that they felt it was their responsibility as a physician to post and/or engage on social media. In what ways, if any, does that resonate with you?
- Did you ever have a patient/patient’s family come into your practice with misconceptions or erroneous medical concepts taken from social media?

**Twitter’s current situation**

Have you been following the discussion around Elon Musk’s takeover of Twitter?

- Has this influenced the way you look at the platform? Would you consider leaving Twitter?
- Do you feel that the takeover has changed the landscape of disinformation on the platform?

**Concluding Questions**

- Is there anything about physicians' use of social media that you think we should be asking about?
- Lastly, we are interested in speaking with additional physicians. When you think of other key physicians that engage with social media, who comes to mind and why?
